# Feasibility, Reproducibility and Cold-Induced Energy Expenditure using Whole-Room Calorimetry in Adults and Children

**DOI:** 10.1101/2025.09.03.25334922

**Authors:** Paige Cheveldayoff, Bader Alamri, Dongdong Wang, Rogelio Cruz Gonzalez, Norm Konyer, Michael D. Noseworthy, Hertzel C. Gerstein, Zubin Punthakee, Gregory R. Steinberg, Katherine M. Morrison

## Abstract

**Context:** To understand energy balance, whole-room indirect calorimetry (WRIC) allows for accurate measurement of energy expenditure (EE).

**Objectives:** To examine the relationship between cold-induced resting EE and brown adipose tissue (BAT) activity measured by MRI, evaluate WRICS performance and feasibility of use in children and adults.

**Methods:** The WRICS was equipped with a Promethion High-Definition Room Calorimetry system. Technical validation utilized N2 and CO2 gas infusions. Healthy adults and children (8 years and older) attended two 4-hour WRIC visits (one week apart) and one MRI visit. Resting EE at 25°C (REE_25_) was compared between visits and to REE at 18°C (REE_18_). Recruitment and completion rates were examined. BAT activity was assessed by MRI as the decline in supraclavicular proton density fat fraction during 18°C cold exposure.

**Results:** Gas infusion testing confirmed high accuracy (RER=0.99; 95% CI 0.991–0.996). Study completion rates were high (Adults: 20/21; Children: 18/18). REE_25_ was consistent between visits (Adults: 1.59 vs 1.63 kcal/min, p=0.56; Children: 1.57 vs 1.56 kcal/min, p=0.76) with good reproducibility (ICC Adults: 0.766; Children: 0.887). Cold exposure increased REE by 0.23 kcal/min (adults) and 0.18 kcal/min (children). BAT activity correlated with REE_18_ in both groups (Adults: r=0.51, p=0.03; Children: r=0.64, p=0.03).

**Conclusion:** WRICS use was feasible in adults and children. The WRICS measurement was accurate, measures of REE were reproducible and changes in EE during cold were measurable, and related to BAT activity, supporting the usefulness of this system in the assessment of EE in response to interventions in adults and children.

## Introduction

Obesity is a common condition that contributes to considerable morbidity and mortality^1, 2^. Energy imbalance resulting from the disparity between energy intake, absorption, and expenditure is a key factor in the development of obesity and its accompanying metabolic complications including metabolic dysfunction-associated steatotic liver disease (MASLD) and type 2 diabetes^3^.

Total energy expenditure (TEE) is comprised of resting energy expenditure (REE), diet- induced thermogenesis (DIT) and physical activity EE which includes both exercise and non- exercise related EE. REE is the amount of energy that the body requires for essential organ and cellular function when lying in a state of physiological and psychological rest. DIT, also known as the thermic effect of food, is energy expended for the digestion, absorption and storage of food. REE accounts for 60% of total EE, while DIT and physical activity EE contribute 10% and 15-30%, respectively^4^. Critically, EE has been noted to decline with weight loss to a greater extent than predicted by changes in body composition alone – a phenomenon termed metabolic adaptation to weight loss or adaptive thermogenesis^5^. Given the potential contribution of metabolic adaptation to weight regain, understanding of the mechanisms contributing to this must be better understood – but requires accurate, reproducible measurement of EE.

It has long been known that energy expenditure can be quantified by measuring respiratory gas exchange, known as indirect calorimetry^6^. Whole room indirect calorimetry allows study participants to move freely in the room and can be used over hours to days, enabling measurement of all components of EE. Furthermore, improvements in technology have enhanced the sensitivity of whole room calorimeters, thereby reducing response times to minutes. To enhance comparability of findings from multiple laboratories, an expert panel established the Room Indirect Calorimetry Operating and Reporting Standards, version 1.0 (RICORS 1.0) in 2019 to establish minimum requirements for reporting technical specifications, data reduction and analytical approaches, performance standards, elements of study design and outcome measures for studies employing whole room indirect calorimetry^7^. These standards will enable comparison of findings relating to human energy metabolism across sites, thereby enhancing knowledge in this important field.

REE is influenced by age, body composition, diet, environmental temperature, biological sex, hormone levels and medications^4, 8–10^. One contributor to variation in REE is brown adipose tissue (BAT) activity through non-shivering thermogenesis^11^. As BAT activity is linked to better metabolic health in adults^12–14^ and in children^15^, there has been a growing interest in understanding factors that contribute to or inhibit BAT activity across the life course. The contribution of changes in BAT activity to metabolic adaptation with weight loss is unknown.

To better understand EE in adults and children, including in response to weight loss, the Centre for Metabolism, Obesity and Diabetes Research at McMaster University has established two new whole room indirect calorimeters. Here we report the technical specifications and performance standards of this new system in the evaluation of REE at 25°C and during an 18°C cold exposure, and DIT in adults and children. We discuss the feasibility of using whole room indirect calorimetry in children, the reproducibility of measurement of REE in children and adults and the ability of the system to detect changes in REE during cold exposure (REE_18_) and after eating a standardized mixed meal. Further, we examine the relationship of cold induced changes in EE and an MRI based measurement of BAT activity in both adults and children.

## Methods

### System Accuracy of the Whole-room indirect calorimetry system

This technical validation complies with the RICORS 1.0 recommendations for initial validation of the two whole room indirect calorimetry systems (WRICS) at the Centre for Metabolism, Obesity and Diabetes Research, McMaster University. The rooms, measuring 3.5m W x 2.9m D x 2.4m H (volume 23.9 m^3^) and 2.3m W x 2.9m D x 2.4m H (volume 15.6 m^3^) (Cantrol Environment Systems Inc, Toronto, Canada), were constructed at McMaster University in Hamilton, Ontario, within the McMaster University Medical Centre. The larger calorimeter, which was used for the current study, is equipped with a sofa bed, chair, table, entertainment station, sink and toilet for participant use. In addition, both rooms are equipped with windows, an intercom for communication, and a dual-lever airtight lock for food or water delivery to participants while in the room.

Respiratory exchange within the chamber, as well as room temperature and humidity were measured using the Promethion GA-3m2/FG-250 model (Promethion High-Definition Room Calorimetry System, Sable Systems International, NV, USA). The manufacturer’s guidelines were adhered to with respect to daily and periodic calibration. CO_2_, O_2_, water vapor pressure, and barometric pressure were measured by the system on a second-by-second basis and subsequently averaged and exported on a minute-by-minute basis by the Promethion system. This data was then averaged over intervals of five minutes. Glucose and fatty acid oxidation rates were calculated with the WRICS using Frayn’s equation^16^. The energy expenditure was determined using the Weir formula^17^.

The system performance portion of the technical validation of the room calorimeter was conducted using the gas infusion method specified in RICORS 1.0^7^. High-purity nitrogen (N_2_) (99.1%) and carbon dioxide (CO_2_) (99.99%) gases (LINDE Canada Inc, Scarborough, Canada) were infused at specific rates using a calibrated mass-flow controller for each respiratory exchange ratio (RER). For an RER of 1.0, CO_2_ was infused at 0.2500 L/min, and N_2_ at 0.9434 L/min. For an RER of 0.8, CO_2_ was infused at 0.2000 L/min, with N_2_ also at 0.9434 L/min. Additionally, in compliance with RICORS 1.0 “zero” or empty room, tests were performed without gas infusion. To ensure the absence of any leaks in the system, standard recovery tests were performed monthly.

### Feasibility and reproducibility of measurement in adults and children

This was a three visit, cross-sectional study of adults and children and was undertaken from October 2022 to August 2024. Participants between the ages of 8 and 40 years who were able to communicate in English were included. Exclusion criteria included a history of type 1 or 2 diabetes mellitus, prior bariatric surgery or liver transplantation, pregnancy or current breast feeding, medications known to influence metabolism, and contradictions for magnetic resonance imaging (MRI) (claustrophobia, implanted metal, metallic injuries, recent tattoo or weight > 136kg). Recruitment occurred through advertisement locally and in the community. Feasibility was evaluated by assessing study recruitment and visit completion, with minimal attrition (<15%) indicating feasibility. The study was approved by the Hamilton Integrated Research Ethics Board (HiREB), and all participants provided either informed consent or a parent/guardian provided informed consent, and the participant provided assent.

Each of the three study visits (two in the larger WRICS and one imaging visit) occurred with the participant fasting for at least 8 hours. The participants were instructed to refrain from ingesting caffeine for at least 12 hours, to refrain from physical activity for 48 hours and to abstain from foods high in serotonin (nuts, avocados, tomatoes, bananas, kiwis, pineapple and plums) for at least 24 hours prior to all study visits, due to potential inhibitory effects on BAT activity^18, 19^. Anthropometrics, body composition and energy expenditure were evaluated at the EE / metabolic visit and questionnaires including demographics and health were completed at the time of the imaging visit. Participants completed a 3-day food log and a questionnaire pertaining to participant demographic information (age, sex, ethnicity, and current medications) and personal/family health history. Pediatric participants completed a self-assessment of pubertal status^20^.

#### Energy Expenditure and Metabolic Testing Visits

The two EE / metabolic visits were conducted one week apart. Prior to EE measurement in the WRICS, anthropometry (height, weight, waist circumference), blood pressure, heart rate and body composition were measured. Height was measured using a wall-mounted stadiometer (Seca, model 24) to the nearest 0.1cm. Weight was measured using a calibrated electronic scale (InBody 570) to the nearest 0.1kg. Waist circumference was measured at the midpoint between the lowest rib and iliac crest using a weighted measuring tape (OHAUS Pull Type Spring Scale, Parsippany, NJ, USA) set at 750g to the nearest 0.1cm. Height, weight and waist circumference were measured three times and their respective averages were calculated. Blood pressure and heart rate were measured following a period of rest where the participants were instructed to sit quietly in an upright supported chair, with their legs uncrossed and to refrain from talking (One Piece Cuff - Suntech, CT40, Morrisville, NC, USA). Blood pressure and heart rate were measured five times for optimal accuracy and then averaged. Body mass index (BMI) was calculated using the averaged heights and weights (kg/m^2^), and, for children, BMI z-score was determined utilizing the standards set by the World Health Organization^21^. Body composition was measured at one visit using dual-energy x-ray absorptiometry (DXA, Lunar PRODIGY Advance 8743; GE, Healthcare, Waukesha, WI). The Mostellar equation (1987)^22^ was used to calculate body surface area (m^2^).

At both WRICS study visits, participants wore a total body water-perfused suit (Med-Eng, Ottawa, Ontario, Canada). Participants were randomized to REE measurement at either 25°C (no water perfusion) or 18°C (suit connected to a flow-controlled circulating bath set at 18°C, located outside the WRICS and with flow-rate of 1L/min, (Isotemp 6200 R28; Fisher Scientific, Waltham, MA, USA). These visits were conducted in a standardized manner (Figure 1). Participants were seated quietly and awake for an initial 15-minute calibration period, and during the following 25-minute period of rest for measurement of REE. To ensure consistent cold exposure, thermocouples (TMQSS-020G-2; OMEGA Engineering, Stamford, CT) fixed to the inlet and outlet manifolds enabled measurement of water temperature and data were recorded and logged at 15 s intervals with a data logger (PowerLab; ADInstruments, Sydney, Australia).

**Figure 1.**
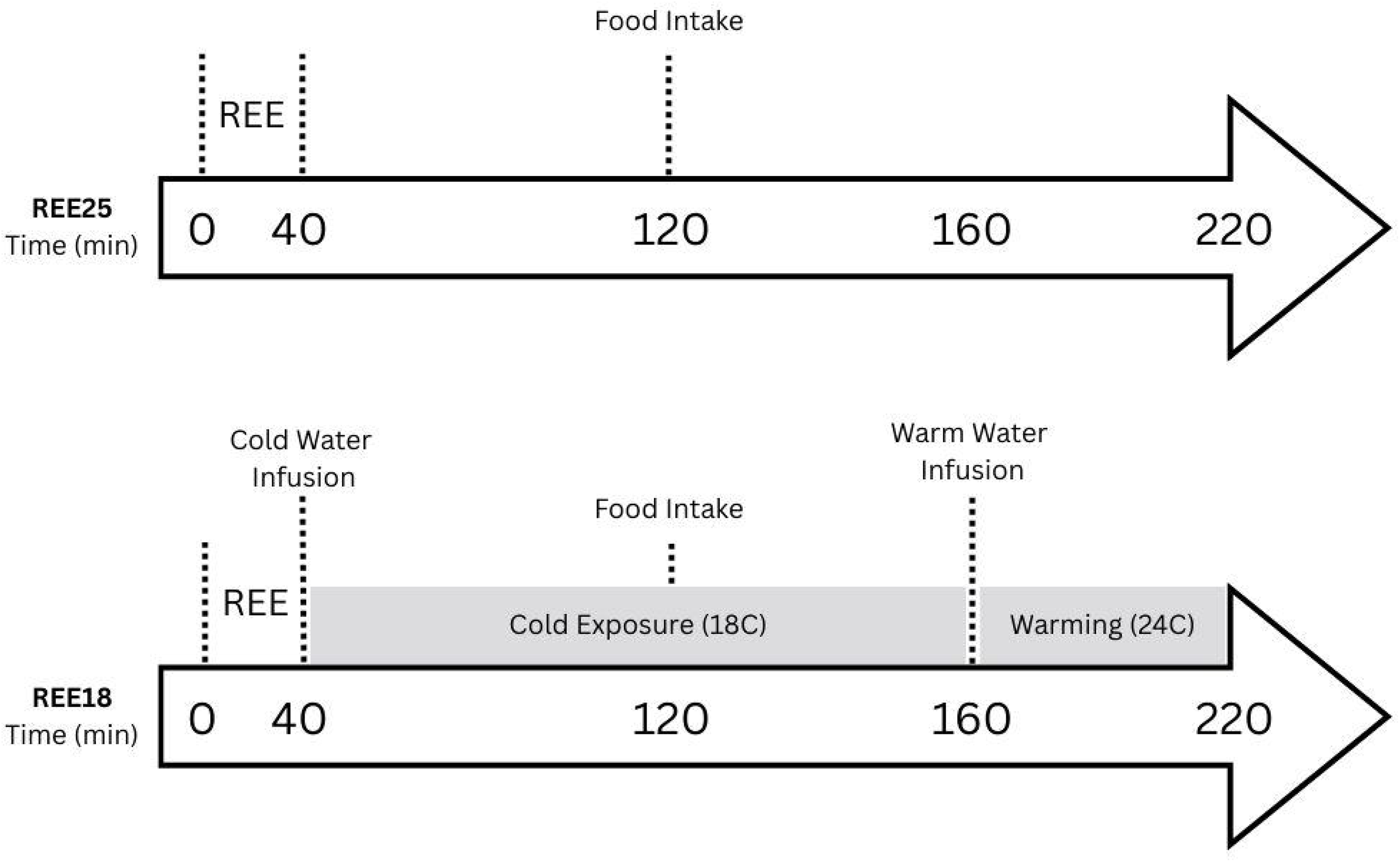
Metabolic visits with cold and at room temperature.

After the two hours of cold exposure, participants underwent a warming period (24°C water) that lasted until the end of their time in the room. A standardized mixed meal comprised of 235ml Ensure Nutrition Supplement (Abbott Nutrition, Chicago, Il, USA) and a 68g CLIF Bar (Mondelez International) (Total: 475 kcal, 74 g carbohydrate, 22 g protein, 12 g fat) was provided at 120 minutes at both visits. REE was measured from minute 15-40, pre-prandial EE, glucose oxidation and fatty acid oxidation from minute 41-120, and post prandial EE, glucose oxidation and fatty acid oxidation from minute 121-240. Following ingestion of the meal, diet induced thermogenesis was measured over a period of two hours and was calculated as the difference between pre-prandial EE and post prandial EE. After 4 hours in the room, participants completed a satiety questionnaire, exited the WRIC and completed post-prandial blood collection

### Imaging visit

#### MRI Studies

One week prior to the first EE/metabolic visit, BAT activity measurement was undertaken at the Imaging Research Centre at St. Joseph’s Healthcare Hamilton (Hamilton, ON, Canada). This involved evaluation of cold-induced changes in the proton density fat fraction in the supraclavicular area with magnetic resonance imaging (MRI) as previously reported^15, 23^.

Briefly, after a 30-minute acclimation period at room temperature, participants underwent the baseline MRI scan with a 3-Tesla whole-body scanner (Discovery MR750; GE Healthcare, Waukesha WI). They were then fitted with the same water perfused suit and flow-controlled circulation bath at 18°C as described above. During the 1-hour cold exposure, the participants lay on a hospital stretcher in a semi-reclined position and were allowed to access their personal or provided devices for movies or music but were not permitted to engage in any movement activities. Measurement of inlet and outlet manifolds was completed as done for EE/metabolic visit, and participants self-reported any shivering. Room temperature and humidity were recorded using a Wireless Forecast Station with Pressure History (model no. WS-9037U-IT; La Crosse Technology, La Crosse, WI).

The MRI protocol and post-imaging analysis were conducted as previously described^15^. Briefly, the proton density fat fraction (PDFF) was obtained using the iterative decomposition of water and fat with echo symmetry and least-squares estimation (IDEAL-IQ) pulse sequence. The PDFF is based on the ratio of the total density of mobile fat protons to the total density of water and mobile fat protons^24^. The PDFF was acquired using a head/neck/spine coil, with an axial image field covering the region between the C2/C3 vertebral disc and the T4/T5 disc (slice thickness 3 mm, 50 slices, flip angle 4°, echo time (TE) 1.3 ms, repetition time (TR) 8.4 ms, field of view (FOV) 340 mm, image resolution 1.52 × 1.42 × 3 mm, acceleration factor 2, and scan time 2.4 min). Six images were generated using this protocol: water-only, fat-only, in-phase, out-of-phase, corrected PDFF, and R2∗ images.

Post-imaging analysis was conducted as previously reported using the AnalyzePro software (version 14; Biomedical Imaging Resource, Mayo Clinic, AnalyzeDirect, Overland Park, KS)^25^. The adipose tissue in the SCV region, between the C5-C6 and T1-T2 disc, was segmented using the sternocleidomastoid, trapezius, and clavicle as medial, posterior, and inferior landmarks, respectively. Regions of interest (ROIs) were drawn manually. A fat-mask and T2 mask (2 to 25 ms) were applied as previously reported – to identify fat tissue (30- 100% fat fraction threshold) and to differentiate BAT from white adipose tissue and muscle. The voxels meeting the above specifications were averaged and reported as SCV PDFF. To correct for partial volume effects, a two-dimensional erosion (1 X 3 voxels) was applied. The percent decline in PDFF (%) is a measure of BAT activity and is calculated as: [(precold SCV PDFF − postcold SCV PDFF)/(precold SCV PDFF)] * 100.

## Statistical analysis

Data analysis was conducted using SPSS statistics (version 29.0) and graphs were constructed using GraphPad Prism 10 (version 10.2). Data are presented as mean ± standard deviation. Descriptive statistics including mean, standard deviation and confidence interval calculations were used to examine the technical validation of the WRICS. Two tailed paired t-tests were used to compare within participant differences in EE and respiratory exchange ratio (RER) and to compare fasting and postprandial fat oxidation and glucose oxidation, and diet-induced thermogenesis between the 25°C and 18°C. One tailed paired t-tests were used to compare fatty acid oxidation and glucose oxidation between the pre and postprandial states. A two-way mixed intraclass correlation coefficient (ICC) was calculated using the variation of interest and total variation for the REE measured at the two visits, approximately one week apart. Pearson correlation coefficient was used to assess the relationship between BAT activity and REE_18_.

## Results

### Technical Validation of the Whole Room Indirect Calorimeters

Using the gas infusion method, the system calculated RER as 0.99 ± 0.001; 95% CI 99.1 – 99.6 for the larger room, and 0.98 ± 0.001; 95% CI 98.0 - 98.5 for the smaller room. When the rate of CO_2_ infusion was changed to 0.2000 standard liters per minute (SLPM) to simulate an RER of 0.8, the calculated RER was 0.82 ± 0.02; 95% CI 78.3 - 86.5, n=7 and 0.809 ± 0.02; 95% CI 80.1 - 81.6, n=4 for the larger and smaller rooms, respectively. To confirm the system’s accuracy, empty room tests with an expected RER of 0.0 were conducted, and the system calculated RER as -0.04 ± 0.07; 95% CI -19.3 – 10.7 for the larger room and -0.04 ± 0.06; 95% CI -18.5– 11.5 for the smaller room. Recovery tests were conducted every month and calibrations were performed every two weeks to ensure the integrity and accuracy of the data collected. These tests involved measuring the amount of CO_2_ present after injecting CO_2_ gas at a rate of 0.2000 L/min overnight. The measured CO_2_ was then divided by the infusion rate and multiplied by 100 to determine the recovery rate.

Recovery rates completed in the larger calorimeter throughout the JOULE study period (ie. November 2022 to August 2024) averaged 93.27± 4.97%; 95% CI 90.6 – 96.9, n=12.

These technical validation results demonstrate the accuracy and reproducibility of the data obtained, indicating that the system possesses a robust structure suitable for conducting human studies. All of the clinical studies described below were conducted using the larger of the two rooms.

### Feasibility and Reproducibility of EE measurement in adults and children

Of the 90 adults and 35 children who expressed interest in the study, 21 adults and 18 children were enrolled (*Figure 2).* One adult participant withdrew before completing the study and has been excluded from analysis.

**Figure 2.**
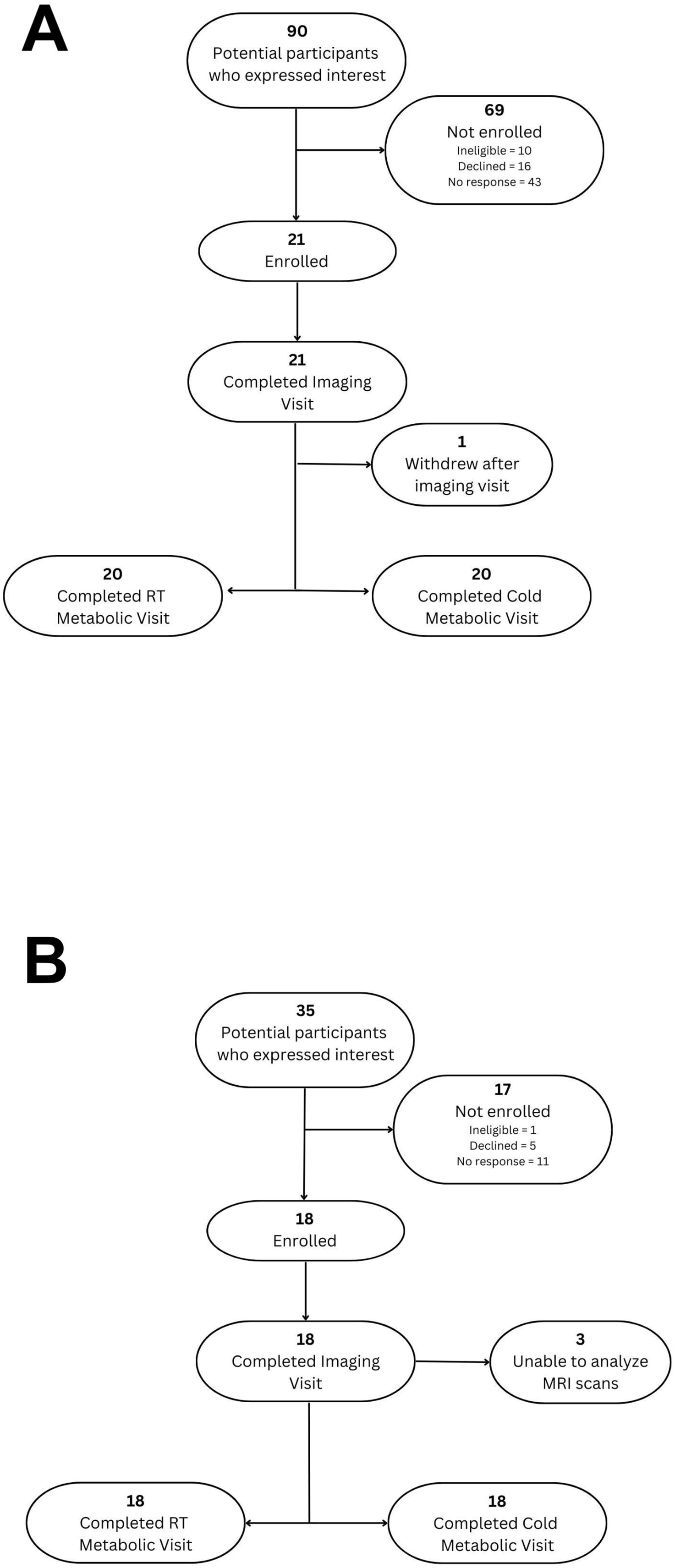
Participant recruitment and study completeness information in 2A: Adults and 2B: Children and adolescents.

The 20 adult (10 female; mean age 24.2 ± 1.2 years) and 18 pediatric (7 female; mean age 13.2 ± 2.8) participants ranged in age from 8-40 years (see Table 1). Three adult participants (all male) had a BMI ≥ 30 kg/ m^2^ and 8 adult participants (3 female, 5 male) had a BMI between 25 and 30 kg/m^2^; thus, 11/20 classified as having overweight or obesity. Amongst the pediatric cohort, 3 participants were classified as having obesity and 2 as having overweight based on WHO BMI z score for age^21^. Four pediatric participants were classified as prepubertal according to the self-assessed puberty staging questionnaire.

**Table 1:** Characteristics of the Study Participants.

| Variable Name (units) | Adult | Pediatric |
| --- | --- | --- |
| Sample Size (n) | 20 | 18 |
| Age (y) | 24.2 ± 5.5 (18, 36) | 13.2 ± 2.8 (8, 17) |
| Sex (M/F) | 10/10 | 11/7 |
| Ethnicity (n) | Caucasian (n=9)<br>South Asian (n=6)<br>Middle Eastern (n=5) | Caucasian (n=15)<br>African (n=2)<br>South Asian (n=1) |
| Tanner puberty stage | N/A | 3.2 ± 1.4 (1, 5) |
| Days between EE visits | 8.25 ± 5.8 (2, 30) | 7.35 ± 5.9 (3, 28) |
| Height (cm) | 169.9 ± 7.8 | 161.7 ± 17.4 |
| Weight (kg) | 74.7 ± 15.9 | 57.4 ± 21.6 |
| Waist circumference (cm) | 61.9 ± 19.3 | 57.4 ± 14.3 |
| Waist/Height Ratio | 0.39 ± 0.06 (0.29, 0.52) | 0.35 ± 0.06 (0.26, 0.47) |
| BMI (kg/m<sup>2</sup>) | 25.6 ± 3.7 (20.2, 32.1) | 21.1 ± 4.9 (15.8, 31.6) |
| BMI z-score (WHO)<sup>21</sup> | N/A | 0.38 ± 0.89 (-0.54, 2.15) |
| Lean Mass Index (kg/m<sup>2</sup>) | 17.52 ± 2.90 (13.2, 22.2) | 14.47 ± 2.19 (10.8, 17.7) |
| Total body fat (%) | 27.8 ± 8.4 (13.6, 45.3) | 26.1 ± 12.2 (6.7, 50.6) |
| Systolic BP (mmHg) | 114.2 ± 9.6 | 107.5 ± 9.6 |
| Diastolic BP (mmHg) | 69.6 ± 7.9 | 66.1 ± 8.2 |
| Heart rate | 68.7 ± 11.5 | 76.3 ± 10.6 |

Table 1: Characteristics of the Study Participants
| Variable Name (units) | Adult | Pediatric |
| --- | --- | --- |
| Sample Size (n) | 20 | 18 |
| Age (y) | 24.2 ± 5.5 (18, 36) | 13.2 ± 2.8 (8, 17) |
| Sex (M/F) | 10/10 | 11/7 |
| Ethnicity (n) | Caucasian (n=9)<br>South Asian (n=6)<br>Middle Eastern (n=5) | Caucasian (n=15)<br>African (n=2)<br>South Asian (n=1) |
| Tanner puberty stage | N/A | 3.2 ± 1.4 (1, 5) |
| Days between EE visits | 8.25 ± 5.8 (2, 30) | 7.35 ± 5.9 (3, 28) |
| Height (cm) | 169.9 ± 7.8 | 161.7 ± 17.4 |
| Weight (kg) | 74.7 ± 15.9 | 57.4 ± 21.6 |
| Waist circumference (cm) | 61.9 ± 19.3 | 57.4 ± 14.3 |
| Waist/Height Ratio | 0.39 ± 0.06 (0.29, 0.52) | 0.35 ± 0.06 (0.26, 0.47) |
| BMI (kg/m<sup>2</sup>) | 25.6 ± 3.7 (20.2, 32.1) | 21.1 ± 4.9 (15.8, 31.6) |
| BMI z-score (WHO)<sup>21</sup> | N/A | 0.38 ± 0.89 (-0.54, 2.15) |
| Lean Mass Index (kg/m<sup>2</sup>) | 17.52 ± 2.90 (13.2, 22.2) | 14.47 ± 2.19 (10.8, 17.7) |
| Total body fat (%) | 27.8 ± 8.4 (13.6, 45.3) | 26.1 ± 12.2 (6.7, 50.6) |
| Systolic BP (mmHg) | 114.2 ± 9.6 | 107.5 ± 9.6 |
| Diastolic BP (mmHg) | 69.6 ± 7.9 | 66.1 ± 8.2 |
| Heart rate | 68.7 ± 11.5 | 76.3 ± 10.6 |

### Reproducibility of REE

REE was measured between 15 and 40 minutes at each of the two visits – prior to the cold exposure. As noted in *Figure 3*, REE was similar between the two visits (Adult: 1.59 ± 0.5, vs 1.63 ± 0.3 kcal/min, Mean ± 95% CI; p-value 0.56; Pediatric: 1.57± 0.34 vs 1.56 ± 0.37 kcal/min, p-value 0.76). The ICC of the REE for these two visits was 0.766 (95% CI [0.408 – 0.908]) for adult participants, and 0.887 (95% CI [0.390-1.74]) for pediatric participants indicating good reliability. Similarly, there were no significant differences in the RER (Adult: 0.83 ± 0.05, 95% CI 0.82 – 0.83 vs. 0.82 ± 0.04, 95% CI 0.82 – 0.83, p= 0.11; Pediatric: 0.84 ± 0.120 vs 0.85 ± 0.014, p= 0.12), room temperature (Adult: 24.6 ± 1.1 °C, 95% CI 24.1-25.1, vs 24.5 ± 1.1 °C, 95% CI 24.0 - 25.0, p = 0.68; Pediatric: 25.4 ± 1.0 °C, 95% CI 24.8 -26.0, vs 24.9 ± 1.2 °C, 95% CI 24.4 - 25.4, p=0.07) or room humidity (Adult: 53.5 ± 5.4%, 95% CI 51.4 - 55.6, vs. 51.7 ± 3.7%, 95% CI 49.6 - 53.8, p = 0.18; Pediatric: 52.3 ± 7.1%, 95% CI 49.4 - 55.4, vs. 52.9 ± 5.3%, 95% CI 49.9 - 55.9, p = 0.73).

**Figure 3:**
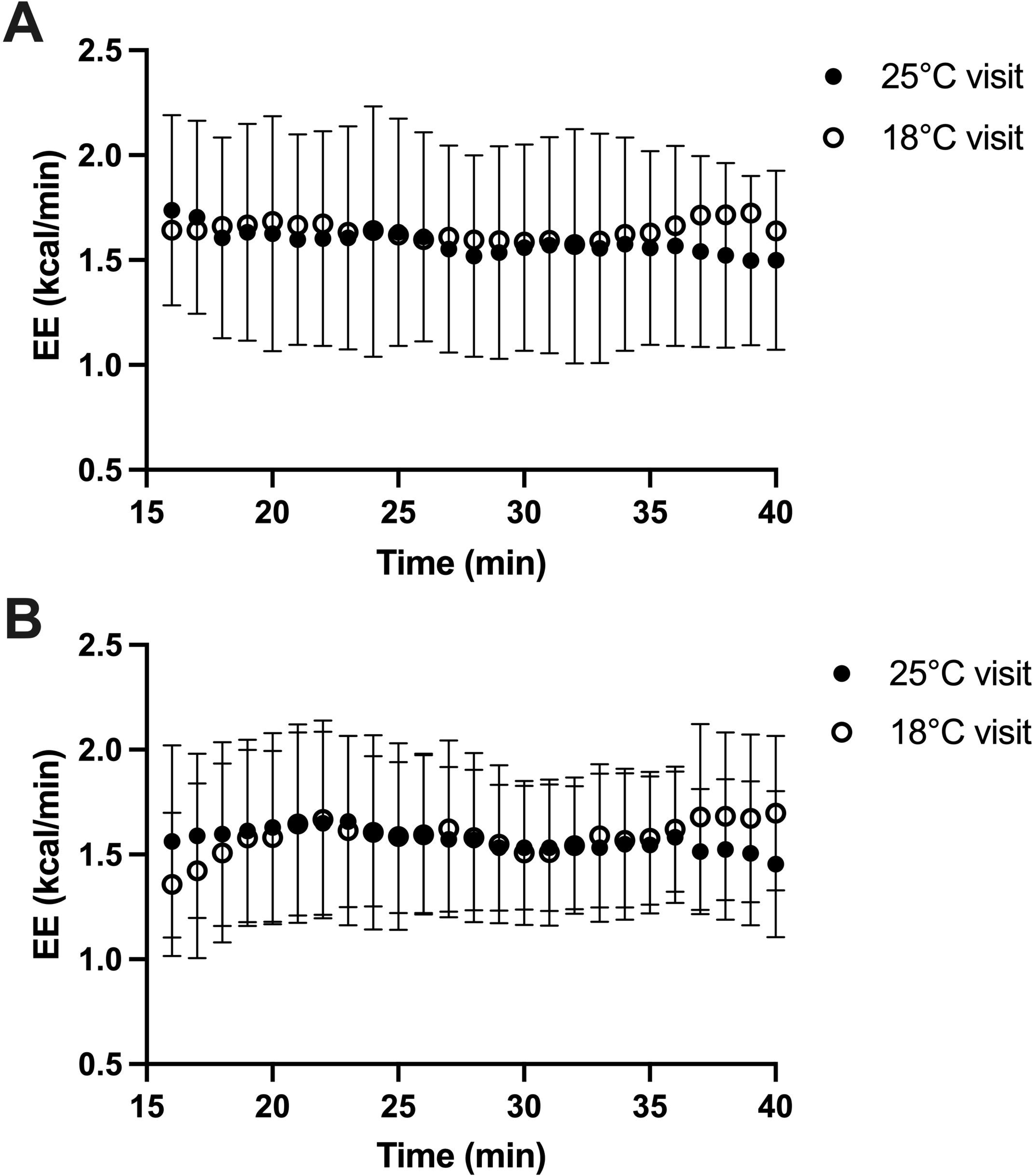
Panel A– Repeatability of REE measures over the two metabolic visits, for adults and B- Repeatability of REE measures over the two metabolic visits, for pediatric participants.

Similarly, there was good reliability of repeat measurements in the smaller room (Adults, n=7; ICC = 0.940 (95% CI [0.760 – 0.991]).

### EE at 18°C or 25°C

After this period at 25°C, participants then either continued at 25°C or experienced a 2-hour period at 18°C (10 adults and 8 children were randomized to experience the cold exposure at their first metabolic / EE visit, the remainder at their second visit).

REE_18_ was higher than REE_25_ in both adults (1.63 ± 0.340 vs. 1.40 ± 0.376 kcal/min, p=0.0013) and children (1.58 ± 0.419 kcal/min vs 1.39 ± 0.320 kcal/min, p = 0.007) (*Figure 4)*. Similarly, postprandial EE_18_ was higher than postprandial EE_25_ in both adults (1.60 ± 0.348 vs. 1.39 ± 0.407 kcal/min; p<0.001) and children (1.62 ± 0.390 vs 1.35± 0.311 kcal/min; p=0.0002). In both age groups and temperature conditions, postprandial EE did not differ from pre-prandial EE. The mean difference between pre- and post-prandial EE in adults was 0.042± 0.19 kcal/min at 18° and 0.032 ± 0.21 kcal/min at 25°C. The mean difference in pre vs post prandial EE in children were 0.044 ± 0.15 kcal/min and 0.042 ± 0.25 kcal/min during the 18° and 25° visits respectively.

**Figure 4:**
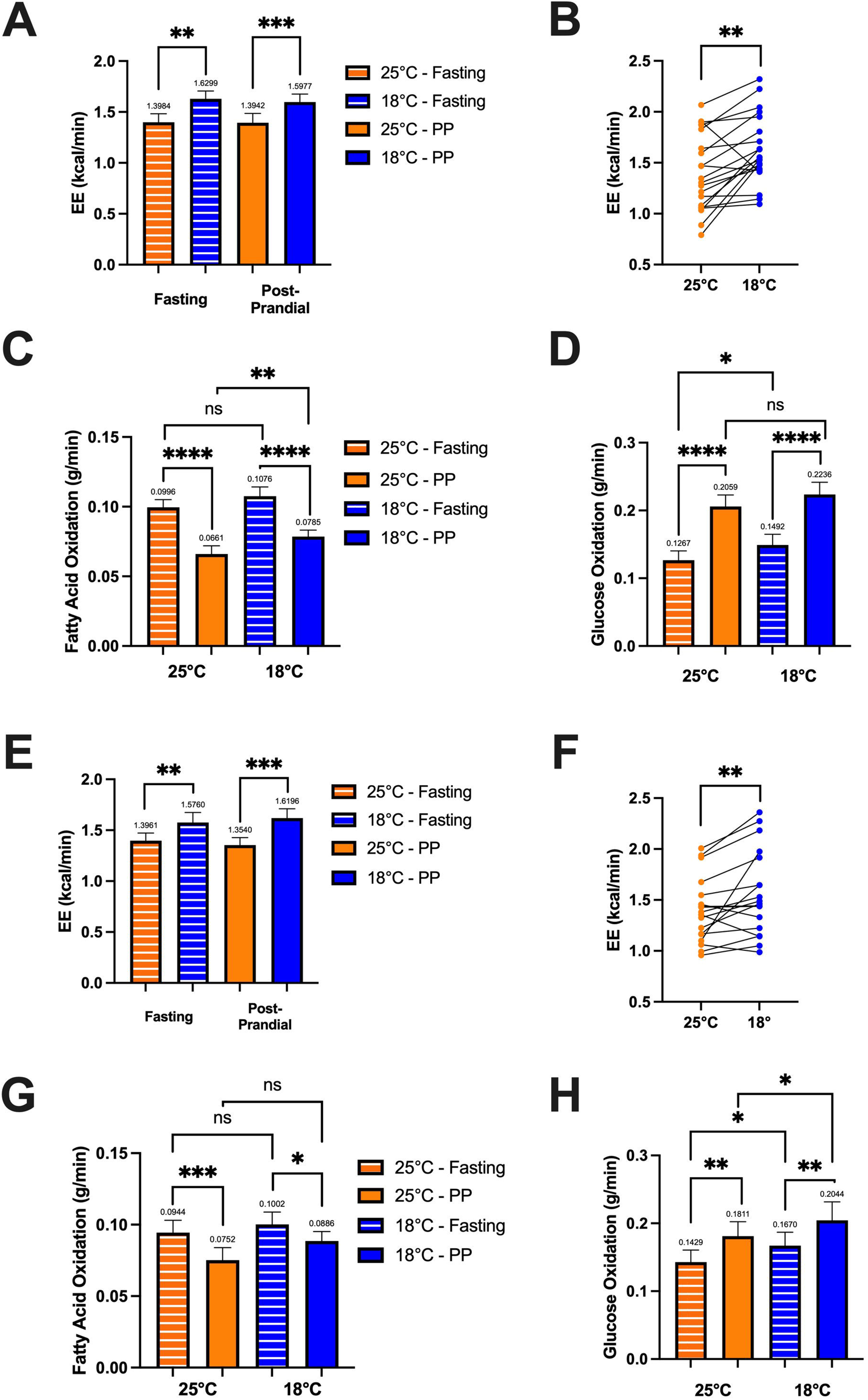
Fasting and postprandial (PP) EE and substrate utilization at 25°C and 18 °C in adults (A-D) and children (E-H). Panel A and B – Fasting EE_18_ greater than EE_25_ (p=0.0013) and post-prandial EE_18_ greater than postprandial EE_25_ (p<0.001). C– Fatty acid oxidation declined postprandially in both 25°C (p<0.0001) and 18°C (p<0.0001) conditions. D – Glucose oxidation increased postprandially in 25°C and 18°C (p<0.0001). Panel E and F – Fasting EE_18_ greater than EE_25_ (p=0.007) and post-prandial EE_18_ greater than postprandial EE_25_ (p=0.0002). G– Fatty acid oxidation declined postprandially in both 25°C (p = 0.0002) and 18°C (p = 0.03) conditions. H –Glucose oxidation increased post-prandially in 25°C (p=0.002) and 18°C (p = 0.004).

### Influence of cold exposure on substrate utilization

In the fasting state, fatty acid oxidation was similar in both the 25°C and cold exposure condition, in both adults and children (Figure 4). As expected, fatty acid oxidation declined after meal intake (Figure 4 –panel C) and the magnitude of decline was similar in the 25°C and cold exposure visits. Glucose oxidation increased after meal intake in adults at both 25°C (0.13 ± 0.062 vs 0.21 ± 0.08 g/min; p<0.0001) and 18°C visits (0.15 ± 0.07 vs 0.22 ± 0.08 g/min; p<0.0001). Similarly, glucose oxidation increased after the meal in the children, both at 25°C (0.14 ± 0.07 vs 0.18 ± 0.09, p =0.002) and 18°C visits (0.17 ± 0.08 vs 0.20 ± 0.11, p = 0.004).

### REE_18_ and BAT activity

The SCV – PDFF declined with cold exposure in 17 of 20 adult participants, suggesting these participants had active BAT tissue (*Figure 5A*) and there were no significant sex differences. Amongst those with evidence of BAT activity, the relative decline in SCV-PDFF with cold was directly related to REE_18_ *(Figure 5A),* but the relationship was not significant when adult participants with no BAT activity were included (p=0.70) Fifteen of eighteen pediatric participants had MRI scans that could be analyzed for SCV- PDFF (three had technical issues preventing SCV-PDFF analysis). A relative decline in the PDFF (mean pre-cold PDFF = 65.99 ± 6.9, mean post-cold PDFF = 64.24 ± 8.5, p = 0.03) was seen, with no differences in the relative decline in PDFF between males 3.9± 6.2 and female 1.7 ± 2.5, p=0.35. Three of the 15 pediatric participants had no decline in PDFF, suggesting no or low BAT activity. As in adults, there was a significant relationship between decline in PDFF and REE_18_ (*Figure 5B*) in BAT active pediatric participants. The relationship was not significant when participants with no BAT activity were included (p = 0.07). In examining the impact of shivering on EE, no significant correlation was found between BAT activity and shivering (Adults, p = 0.32; Children, p= 0.37). Furthermore, no significant differences were found in EE for participants who reported shivering during cold exposure compared to those that did not.

**Figure 5.**
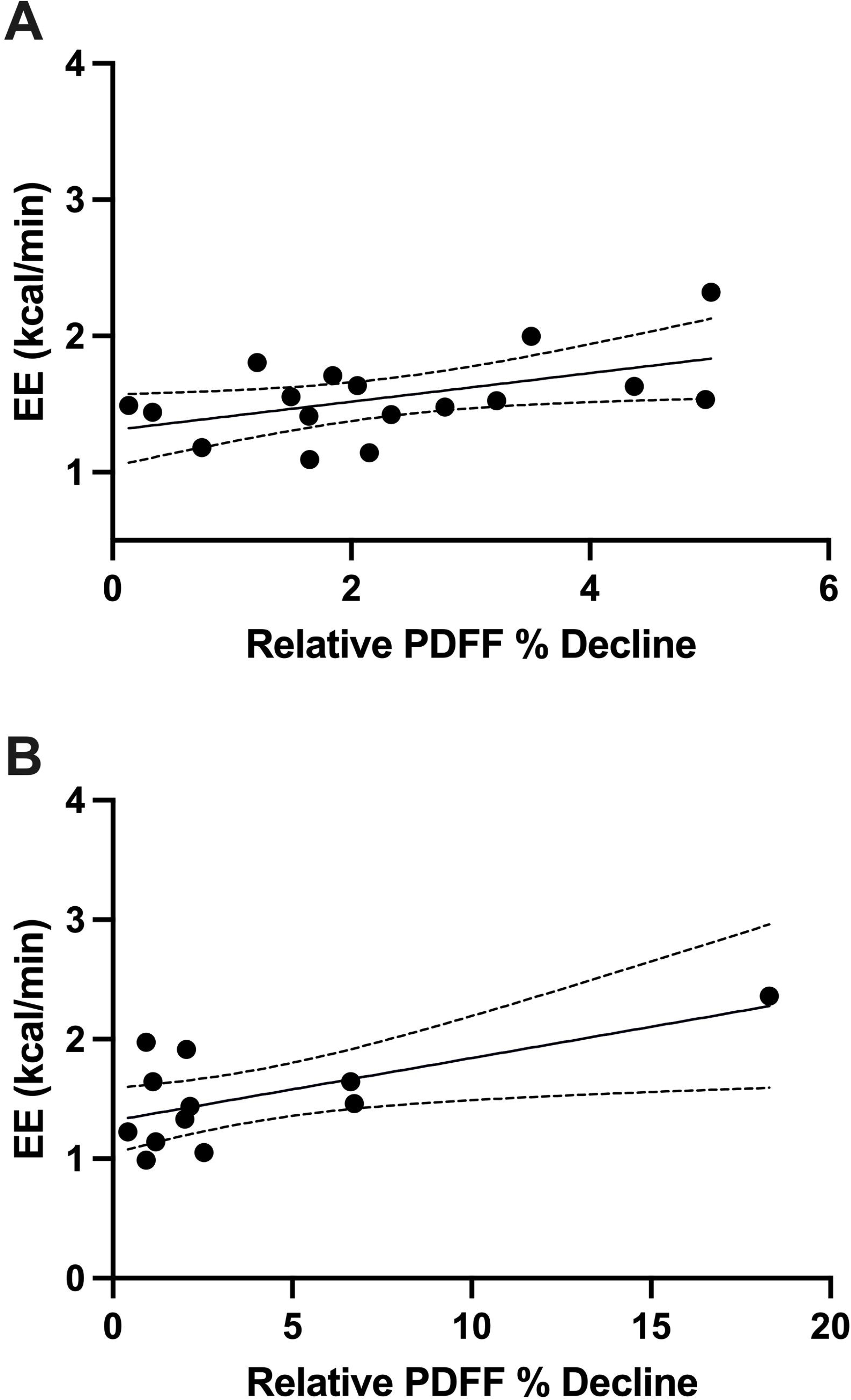
Panel A: Correlation between REE and decline in PDFF among BAT active (n=17) adult participants (r = 0.51, p = 0.03). Panel B: Correlation between REE and decline in PDFF among BAT active (n=12) pediatric participants (r = 0.64, p = 0.03).

## Discussion

The aim of the current study was to evaluate the relationship between cold-induced resting EE and BAT activity using a newly established WRIC system, and to explore the feasibility, accuracy and reliability of this system to document changes in EE during cold exposure and after eating a standardized meal in both adults and children. Following the recommendations of the RICORS 1.0 standards for technical and human validation, this study demonstrates the WRICS system is technically valid, enables reproducible measurement of EE over a brief period (25 min), and on repeated assessments conducted one week apart, can detect a 9-15% increase in EE with cold exposure and it is feasible to have children as young as 8 years of age in the WRICS for up to 4 hours.

Technical validation of the WRIC was undertaken, adhering to the RICORS 1.0 recommendations. The gas infusion method was utilized during dynamic performance measurement for safety reasons and as it offers greater flexibility in adjusting the infusion rate to alter necessary parameters, an advantage not provided by the gas combustion technique^26^. Previous research studies that have utilized both gas infusion and combustion concluded that gas infusion is a sufficient method^27^. The results of the recovery and RER tests using the gas infusion tests were as expected and consistent with the current literature^27–29^. The results of the recovery and RER tests using the gas infusion tests were as expected and consistent with the current literature.

In evaluating feasibility of conducting studies in humans as young as 8 years of age, we note the high completion rate in both adults (95%) and children (100%). Although the feasibility of use of a WRIC in adults was similar to previous studies with completion rates of 95- 98%^28,30,31^, completion of studies in 100% of children in the present study exceeds many previous studies of children 7 – 18 years where completion varied from 74- 96%^32–34^.

The shorter resting period (25 minutes) and time in the WRIC (4 hours) of this study may have positively impacted the recruitment and completion rates and has the potential to aid researchers in completing pragmatic trials with accurate measurements of EE in children. Previous studies have measured REE in children over periods of 1-8 hours^5, 33, 35^. Our experience of reproducible results when REE was measured over a period of only 25 minutes is consistent with a recent study in adults by Rising et al which demonstrated that using 30 versus 60-minutes of WRIC data to estimate REE and RER led to no differences and very high correlation with no bias^36^. Thus, the resting period used in the present study can provide accurate and valid REE measurement without compromising participant comfort and study compliance. Our finding that, measured one week apart, the REE measurement was reliable, with an ICC of 76.6% in adults and 88.7% in children, supports the use of shortened protocols in longitudinal studies as well.

The responsiveness of the system is further demonstrated in its ability to measure changes in EE in response to mild cold exposure. Cold exposure at 18°C for 2 hours resulted in 0.23 kcal/min (15%) higher EE in adults and 0.19 kcal/min (9%) higher EE in children. These results are coherent with a recent meta-analysis which included 10 studies in healthy adults (total n = 171, aged 20-40 years, BMI 20-34 kg/m^2^) that demonstrated an average increase in EE of 188.43 kcal/day (0.13 kcal/min) during cold exposure (varying from 16-19°C) compared to room temperature^37^. Of these 10 studies, the study by Brychta et al. (2019) included the most similar methodology to the present study including whole-room indirect calorimeter use, 30-minute REE measurement, room temperature at 23-25°C, and 16°C air temperature for the cold exposure^38^. These authors report a slightly lower increase in EE compared to the present study with a 0.23 kcal/min increase; however, this may have resulted from study participant and the methodological differences, including the medium of exposure^38–40^. As well, the results of the current study are in line with estimated predictions from energy cost models based on human heat loss with radiation and convection which predict an increase in REE of 0.1 - 0.4 kcal/min with cold^40, 41^. Increased EE with cold results from increases in both shivering and non-shivering thermogenesis^11^. Shivering thermogenesis, characterized by involuntary muscle contractions, elevates metabolic rate and generates heat^42^ whereas non-shivering thermogenesis involves the activation of BAT^43^. We identified no difference in the cold induced increase in EE between those who did and did not report shivering, although shivering was not directly measured with EMG. We did identify a direct relationship between BAT activity and REE during cold in both adults and children.

This result is consistent with previous work in adults undertaken by Blondin et al (2015)^44^ and Tay et al (2020)^45^. The former study identified a positive relationship between BAT activity and EE_18_ as REE increased 1.8-fold following a 180-minute 18°C cold exposure period using a water perfusion suit, compared to that at room temperature^44^. This increase in EE with cold was greater than that of the current study, however, the length of the cold exposure period was also 1 hour longer. In the latter study, researchers denoted a positive relationship between BAT activity and EE_18_ as REE increased by 13% following a 45-minute exposure to a room at 18°C^45^. In another similar study that examined BAT activity and EE before and after cold exposure in healthy adults, researchers reported that BAT activity is the major determinant of EE following cold exposure as only the BAT active individuals demonstrated significantly increased EE in response to cold^46^. In contrast to the findings in response to cold exposure – no increases in EE after eating a standardized meal were demonstrated in either adults or children in the 25°C visit. The 475-kcal meal was given to all participants, regardless of body size and this may have influenced our findings (ie. insufficient caloric load in larger participants). Other studies have provided a meal with an energy intake based on a standardized percentage of estimated required daily energy intake^47^, or as a proportion of measured resting energy expenditure^48^. Given the variability in body size of our participants, the uniform caloric load used in the present study, regardless of individual energy requirements, may have contributed to the blunted thermogenic response observed in both adults and children. Although our DIT measurement of ∼ 0.043 kcal / min is lower than expected based on findings of a systematic review of studies in adults in which DIT was estimated to increase by 1.1 kJ/hour (0.004 kcal/min) for every 100kJ (24 kcal) higher intake, DIT is also altered by meal composition, especially protein content, which varied in the review^49–52^. Although we did not detect an increase in EE it is important to note that the WRICS did detect a drop in RER and the resultant increase in glucose oxidation and reduction in fatty acid oxidation consistent with the composition of the meal.

It is important to note that the adults and children were not fully reclined or fully resting with eyes closed while REE was being measured. While this has the potential to result in a higher REE, the present study reported mean values of 1.5 - 1.6 kcal/min in adults and 1.5 kcal/min in children, which is consistent with findings reported in the current literature of 1.2-1.7 kcal/min for REE in adults and children^30,31^. In addition, it is reassuring that measures were reproducible on two different occasions, even without this strict limitation on cognitive activity.

In conclusion, in addition to documenting the technical validity of our system, the feasibility of measuring EE in children as young as 8 years of age and the repeatability of REE over a relatively brief period we have demonstrated increased EE with cold exposure and the magnitude of this increase is related to BAT activity. These findings provide a strong foundation for future longitudinal, interventional clinical studies.

## Data Availability

Some or all datasets generated during and/or analyzed during the current study are not publicly available but are available from the corresponding author on reasonable request.

## Author Contributions

Conceptualization was done by K.M.M and G.R.S. Methodology was performed by P.C, B.A and R.C-G. Data collection and curation were done by P.C, B.A and R.C-G. Analysis was completed by P.C and B.A. Original manuscript was written by P.C. and B.A. Review and editing were performed by P.C, B.A, D.W, R.C-G, N.K, M.D.N, H.C.G, Z.P, G.R.S, and K.M.M.

---

Supplementary Figure 1- Panel A: Correlation between REE and decline in PDFF among all adult participants (n=20) (r = 0.09, p = 0.70), and B: pediatric participants (n=15) (r = 0.48, p = 0.07).

Supplementary Table 1 – List of Excluded Medications

